# Persistent Cytotoxic Immune Signaling in Anti-VEGF–Treated Neovascular Age-Related Macular Degeneration

**DOI:** 10.64898/2026.04.06.26350115

**Authors:** Marcus A. Toral, Benjamin Ng, Gabriel Velez, Jing Yang, Stephen H. Tsang, Alexander G. Bassuk, Vinit B. Mahajan

## Abstract

**Purpose:** Anti-vascular endothelial growth factor (anti-VEGF) therapy is the standard of care for neovascular age-related macular degeneration (AMD), yet many patients exhibit persistent retinal degeneration, fibrosis, and incomplete therapeutic response. The molecular pathways underlying this incomplete response remain poorly understood. We sought to identify VEGF-independent signaling pathways active in the vitreous of anti-VEGF–treated AMD patients.

**Methods:** We performed multiplex antibody-based proteomic profiling of 1,000 human proteins in vitreous samples from patients with neovascular AMD receiving anti-VEGF therapy (n=8) and comparative controls (n=6). Differential protein expression was assessed using one-way ANOVA, followed by gene ontology and pathway enrichment analyses. Drug–target relationships were evaluated to identify potential opportunities for therapeutic repositioning.

**Results:** We identified 107 differentially expressed proteins (p<0.05), including key regulators of immune signaling, angiogenesis, and metabolism. Notably, multiple components of cytotoxic lymphocyte pathways were dysregulated, including IL-21R, SIGLEC-7, CTLA4, and IL-2–associated signaling. Enrichment analyses revealed significant activation of pathways related to T-cell activation, interleukin signaling, and leukocyte-mediated cytotoxicity. These immune signatures persisted despite suppression of VEGF signaling. Several clinically available immunomodulatory agents—including abatacept, sirolimus, and dupilumab—targeted pathways identified in this dataset.

**Conclusions:** Anti-VEGF–treated neovascular AMD exhibits persistent cytotoxic immune signaling in the vitreous, suggesting that VEGF-independent immune mechanisms may contribute to ongoing retinal damage and incomplete therapeutic response. These findings provide a rationale for combination therapeutic strategies targeting both angiogenic and immune pathways in AMD.

## INTRODUCTION

Age-related Macular Degeneration (AMD) is a chronic inflammatory disease of the retina, and although anti-VEGF therapy is effective at limiting neovascularization, there is persistent fibrosis, cellular atrophy, retinal degeneration, and edema.^1,2,3,4,5^ Recent evidence also indicates that anti-VEGF therapy may contribute to the progression of exudative AMD to the “dry” geographic atrophy form of the disease.^4,6,7^ While the importance of the VEGF-A molecule in driving exudative AMD pathology has become clear, the molecular signals which persist or intensify throughout anti-VEGF therapy are unknown. The success of targeting a single molecule to treat this disease indicates that there may be additional molecules which may be targeted to improve therapy and address persistent pathological features.

We previously found that patients treated with anti-VEGF (bevacizumab) showed increased levels of other angiogenic biomarkers (such as VEGF-C, angiopoietin 2, endothelin 1, follistatin, HB-EGF, HGF, and interleukin 8) despite reduction in VEGF-A levels.^8^ However, this study only examined nineteen specific protein biomarkers for angiogenesis. Additionally, this study tested fluid from the anterior compartment of the eye for analysis. While this region of the eye is easier to access surgically, it is unclear whether it is representative of the posterior compartment of the eye, where the primary disease manifestations occur in the context of AMD.

Here we sought to build upon our previous work, performing large-scale proteomic analyses of vitreous humor biopsies from patients with exudative AMD during anti-VEGF treatment with bevacizumab. Proteomic analysis, the large-scale study of proteins, is a powerful tool for identifying new biomarkers and therapeutic targets in human disease. Using this approach, we have found that undiluted, vitreous biopsies are a good source of biomarkers that indicate retinal health.^9,10^ While human vitreous tissue is exceptionally difficult to obtain, particularly from AMD patients (who are typically not good surgical candidates), a relatively small sample size is appropriate for the identification of candidate biomarkers in the discovery stage of a rigorous proteomic workflow.^9,10,11,12,13,14^

To date, there are only a handful of proteomic analyses using human AMD tissue, and there is limited consistency among their findings (Table 1). The discrepancies between these studies may be partially explained by the predominant reliance on mass spectrometry-based approaches during the biomarker discovery phase. While the approach is powerful, it remains difficult to accurately quantify data from liquid chromatography coupled with tandem mass-spectrometry (LC-MS/MS) experiments. In LC-MS/MS, highly abundant protein signals can drown out less abundant (but often biologically-relevant) proteins, and depletion of abundant proteins (e.g., albumin) prior to analysis may remove many less-abundant protein interaction partners (e.g., proteins which bind albumin).^15^ In contrast, antibody array panels can readily quantify protein targets independent of non-target protein levels, and experimental evidence supports this performance advantage.^11^ To date, no proteomic analysis of living exudative AMD patient vitreous has employed a large multiplex antibody-based approach. Here, we used a high-throughput antibody array to detect and quantify 1,000 different protein biomarkers in vitreous biopsies from AMD patients following anti-VEGF treatment.

**Table 1.** Summary of AMD Vitreous Proteomic Analyses.

| Paper | Proteomics Platform | Highlighted upregulated proteins | Highlighted downregulated proteins |
| --- | --- | --- | --- |
| Toral, 2019<br>(Present study) | Multiplex antibody microarray | SIGLEC-7<br>PCK1<br>SPHK1<br>ST3GAL1 | IL-21R<br>CDH6<br>CTLA4<br>SULT2A1 |
| Schori, 2018[42] | LC-MS/MS | VEGFR1<br>1433G<br>AL1A1<br>FRIH | PROCR<br>BLMH<br>ASPRV1<br>SPTA1 |
| Nobl, 2016[43] | CE-MS<br>LC-MS/MS | Prostaglandin-H2 alpha-isomerase<br>Ig gamma-1 chain C region<br>Ig lambda-2 chain C regions<br>Serotransferrin | Collagen alpha-1 (VII) chain<br>Opticin<br>Alpha-crystallin A chain<br>Obscurin |
| Koss, 2014[44] | CE-MS<br>LC-MS/MS | Ig alpha-1 chain C region<br>Histidine-rich glycoprotein<br>Ig kappa/lambda chain C region<br>Anti-thrombin III | No data available |
| Ecker, 2012[45] | Reverse phase protein microarray | MMP-9<br>IL-12<br>cABL Thr735<br>Heme oxygenase-1 | No data available |

\*Abbreviations: LC-MS/MS = Liquid chromatography-mass Spectrometry / mass spectrometry. CE-MS = Capillary electrophoresis mass spectrometry.

## METHODS

### Study Approval

The study design and protocol received approval from the Stanford Institutional Review Board (IRB; protocol #42754). Written informed consent, in accordance with the tenets of the Declaration of Helsinki for Human Research and the Health Insurance Portability and Accountability Act (HIPAA), was obtained from all participants. Vitreous samples were collected and analyzed between August 2011 to January 2016.

### Vitreous Sample Collection

Pars plana vitrectomy was performed using a single-step transconjunctival 23-gauge trocar cannular system (Alcon Laboratories Inc, Fort Worth, TX), and an undiluted 1-cc sample of the vitreous was manually aspirated into a 3-cc syringe.^16,17^ Vitreous samples were collected from eight patients with exudative AMD following recent bevacizumab anti-VEGF treatment, and six comparative control patients with idiopathic non-inflammatory epiretinal membranes (Table 2). Specific anti-coagulation measures were not used with vitreous samples. To address the possibility of blood contamination as a confounding variable in our data, we also collected five vitreous samples from non-diabetic patients with vitreous hemorrhage (VH) and an additional five idiopathic epiretinal membrane patients for comparison. Vitreous samples were immediately centrifuged in the operating room at 15,000 x g for 5 minutes at room temperature to remove impurities and then stored at -80°C, as previously described.^18^

**Table 2.**
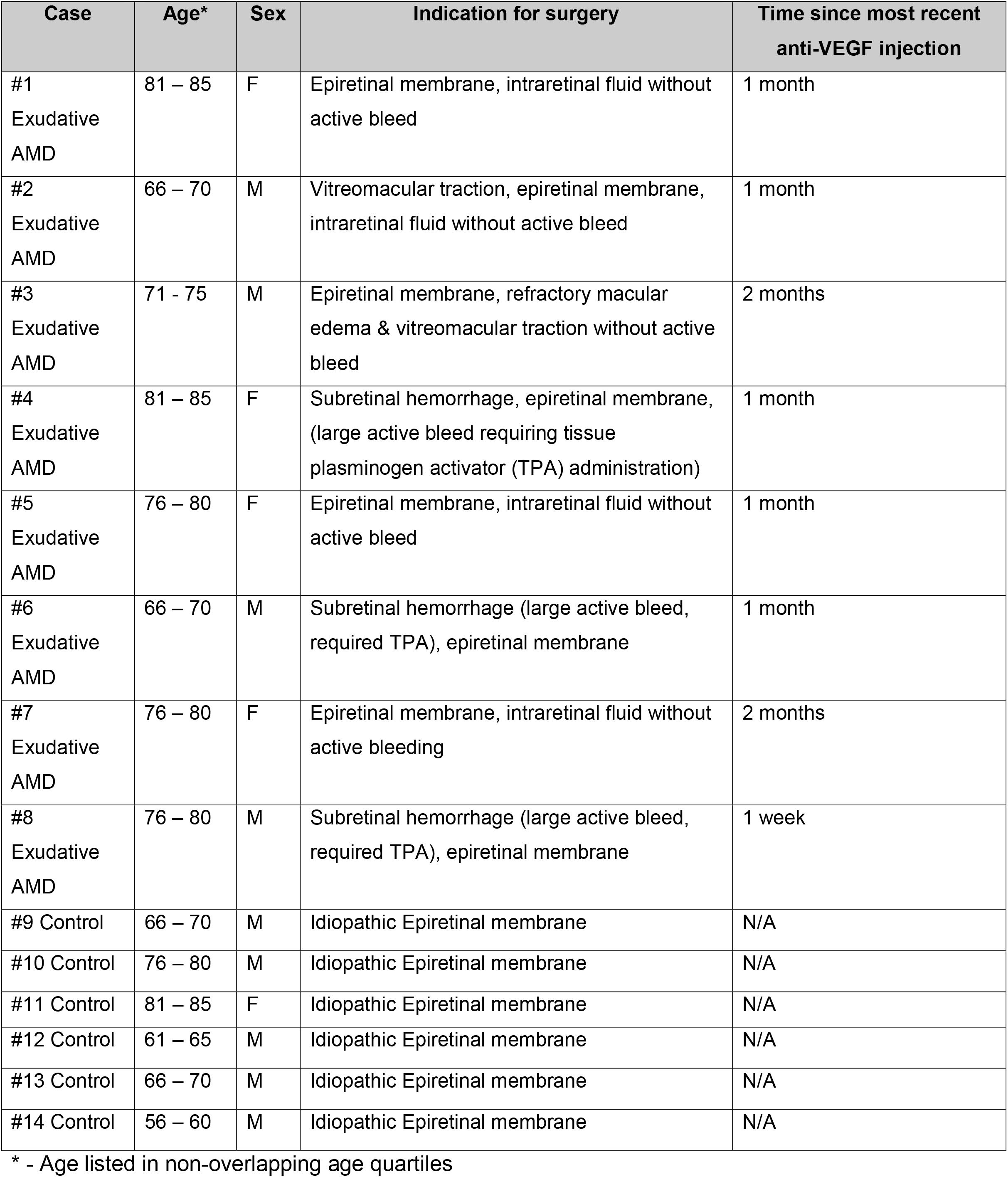
Patient Demographics and Details.

### Protein Biomarker Multiplex Array

Vitreous proteins were measured using the Human Quantibody Kiloplex Array (RayBiotech, Norcross, GA) per the manufacturers protocol. This array detects 1,000 human biomarkers. First, the array chips were incubated with sample diluents for 30 minutes at room temperature to act as a block. Diluted vitreous (four technical replicates per sample; 1.5 mL) was added to the wells of the array and incubated overnight at 4°C. A standard protein dilution was added to the wells of the array to determine protein concentrations. For signal detection, 80 µL of Cy3-Streptavidin was added to each well, rinsed and visualized by laser scanner. The RayBio^®^ Analysis Tool (RayBio^®^, Norcross, GA) was used for protein classification. Final protein concentrations (in pg/mL) were corrected for sample dilution.

### Statistical and bioinformatics analysis

Protein concentrations were measured, and data were sorted on Microsoft excel spreadsheets based on concentration hierarchy. Data were sorted into two sections: comparative control patients with idiopathic epiretinal membranes, and patients with exudative AMD (as well as, separately, VH and control sections to rule out blood contamination). Results were saved in Excel as .txt format and were uploaded into the Partek Genomics Suite 6.5 software package. A strict fold-change cutoff was not employed.^19^ The data for control and exudative AMD patients were normalized to log base 2 and compared using one-way ANOVA. While a fold-change cutoff was not used, a significance threshold was applied for consistency with previous studies.^12,13,20^ All proteins with non-significant (p ≥ 0.05) changes were eliminated from the table. The significant values were used to generate a heatmap using the ‘cluster based on significant genes’ visualization function with the standardization option.

### Gene ontology and pathway analysis

For gene ontology analysis, two lists were uploaded onto the WebGestalt^21^ online bioinformatics toolkit: (1) A list of the 1,000 protein biomarkers present on our array, and (2) A list of significantly differentially expressed proteins (p<0.05). Overrepresentation Enrichment Analysis (ORA) was carried out using the WebGestalt gene ontology functional database (enrichment category: Biological Processes). Similarly, PANTHER^22^ protein pathway analysis was performed using the WebGestalt tool.

## RESULTS

### Patient cohort demographics

Vitreous biopsies were collected from eight exudative AMD patients and six comparative control patients (Table 2). AMD patients had been treated with bevacizumab injections within the past one week to two months. Three had large subretinal hemorrhages that underwent subretinal tissue plasminogen activator (TPA) injection. Vitreous was subsequently analyzed on multiplex antibody arrays as outlined in the methods. Since evidence of subretinal bleeding was observed in three of eight of AMD samples, but none of the controls, vitreous biopsies were also collected from five non-diabetic patients with VH (Supplemental Table 1). These samples served as a comparative control for possible vitreous blood contamination, allowing us to identify potential confounders from the systemic blood circulation in our dataset not unique to AMD.

### Statistical analysis and hierarchical clustering reveal differentially expressed proteins

Principal component analysis revealed two distinct clusters corresponding to the two experimental groups: exudative AMD and controls (Figure 1A). One-way ANOVA and hierarchical heatmap clustering identified 107 differentially expressed proteins (p < 0.05; Figure 1B). Sixty-two proteins were significantly upregulated in AMD compared to controls, while 45 were downregulated. AMD samples with and without bleeding clustered together on both PCA and hierarchical heatmap clustering, indicating blood contamination was likely insignificant. This conclusion was further supported experimentally by analysis of VH samples, which demonstrated a molecular signature distinct from AMD and prominently featured protein pathways consistent with blood contamination which were not identified in the AMD cohort (Supplemental Figure 1, Supplemental Data 1&2).

**Figure 1.**
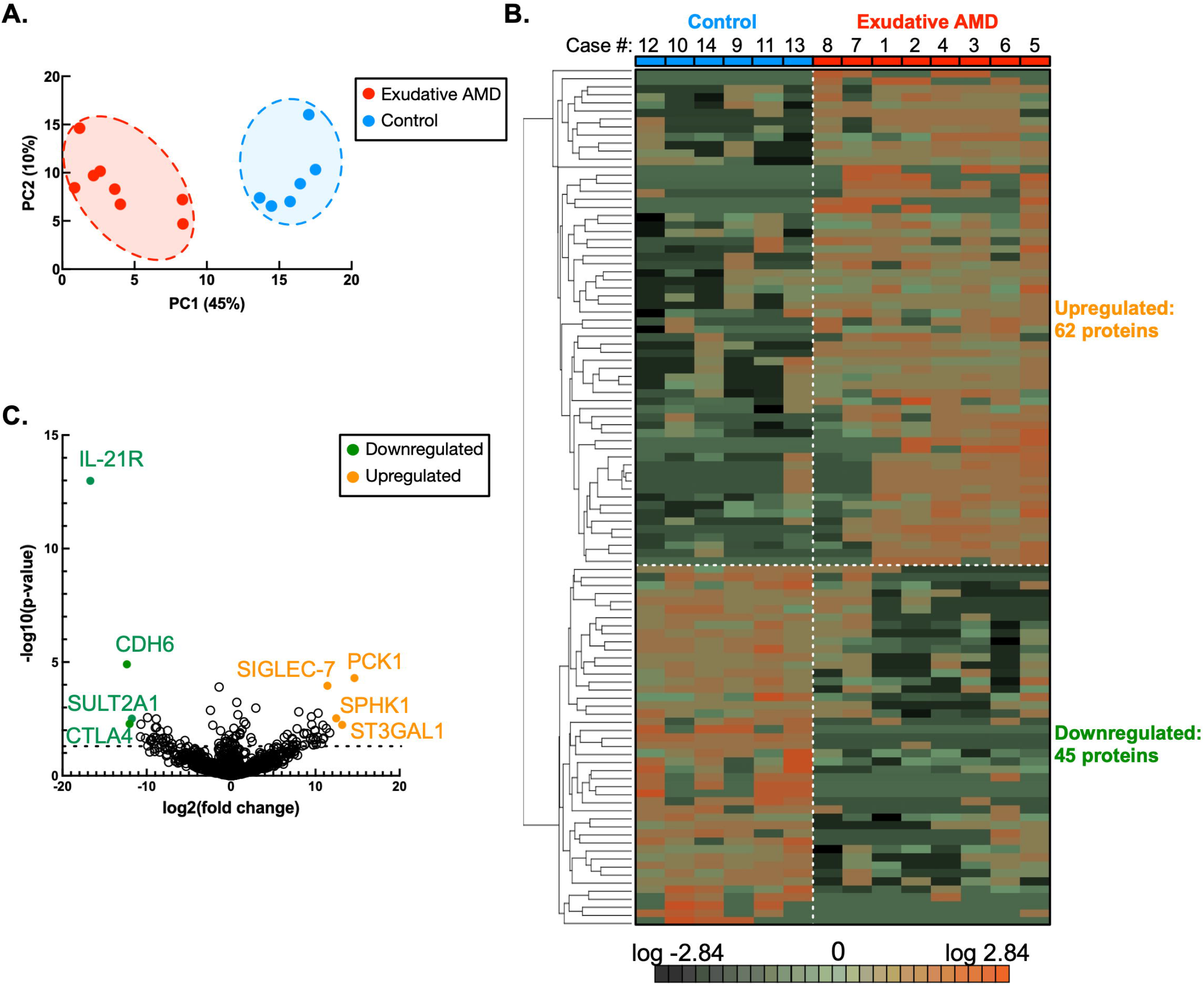
Proteomic analysis identifies unique exudative AMD molecular signature. **A)** Protein levels (pg/mL) from the multiplex ELISA assay were analyzed by one-way ANOVA and principal component analysis (PCA). One-way ANOVA identified 95 proteins that best differentiate AMD vitreous from control vitreous. The score plot of PC1 and PC2 showed separation between eight AMD samples (red) and six control samples (blue) based on the levels of these 95 proteins. **B)** Protein concentrations from the multiplex ELISA array were log2 normalized and analyzed for differentially expressed proteins. Multi-group comparison (one-way ANOVA) followed by hierarchical heat map clustering was used to identify differentially expressed proteins in the large-scale dataset. The analysis detected 107 proteins with statistically significant (p<0.05) expression between AMD and control samples. The results are represented as a heatmap and display protein expression levels (fold change) on a logarithmic scale. Orange indicates high expression (62 proteins) while dark green/black indicate low or no expression (45 proteins). **C)** Differentially expressed proteins were represented as a volcano plot. The horizontal axis (x-axis) displays the log2 fold-change value (AMD vs controls) and the vertical axis (y-axis) displays the noise-adjusted signal as the -log10 p-value obtained from the one-way ANOVA analysis. Proteins emerging from the plot with particularly small p-values and large fold changes which we focused on in this analysis are highlighted in green (downregulated) and orange (upregulated). The proteins PCK1, SIGLEC-7, ST3GAL1, and SPHK1 were highlighted as upregulated in AMD samples, while IL-21R, CDH6, SULT2A1, and CTLA4 were highlighted as downregulated in AMD samples.

In AMD samples, our analysis revealed that the proteins interleukin-21 receptor (IL-21R), sialic acid binding Ig like lectin 7 (SIGLEC-7), cytotoxic T lymphocyte associated protein 4 (CTLA4), phosphoenolpyruvate carboxykinase 1 (PCK1), sphingosine kinase 1 (SPHK1), ST3 beta-galactosidease alpha-2,3-sialytransferase 1 (ST3GAL1), cadherin-6 (CDH6), and sulfotransferase family 2A member 1 (SULT2A1) showed particularly significant expression changes (Figure 1C). The differentially expressed proteins were diverse but implicated specific signaling pathways. Altered levels of IL-21R, SIGLEC-7, SIGLEC-9, ST3GAL1, interferon gamma (IFNγ), CTLA4, and interleukin-2 (IL-2) indicated disrupted signaling relevant to subclasses of natural killer cells and cytotoxic T-cells. Separately, SULT2A1 was significantly downregulated. SULT2A1 is a sulfotransferase possibly involved in modest processing of 7-ketocholesterol, known to play a role in AMD.^23^ CDH6 was also highly downregulated and is thought to be involved in retinal development, although its role in adult vitreoretinal tissue is unclear.^24^

As expected, there were low levels of VEGF without a significant difference between the two groups, consistent with prior anti-VEGF treatment. Nonetheless, we detected significant upregulation of several other angiogenic biomarkers, including PDGFRβ, IGF-1, and GROα. SPHK1, an enzyme involved in regulating angiogenesis and neuroinflammation, was also upregulated. Additionally, there was upregulation of PCK1, the rate-limiting enzyme of gluconeogenesis. Indeed, PCK1 dysregulation has been implicated in angiogenesis in cancer, and disruption of glucose metabolism may play a role in driving choroidal neovascularization.^25^

### Gene ontology and protein pathway analysis indicate active molecular pathways

We next performed gene ontological (GO) analysis to identify the top highly enriched protein categories (Figure 2A). The top 5 enriched biological process categories were: drug catabolic process, antibiotic catabolic process, ciliary neurotrophic factor-mediated signaling pathway, embryonic hemopoiesis, and retina homeostasis. Additionally, we observed enrichment of GO categories for regulation of axon regeneration and regulation of neuron projection regeneration, consistent with the damaged neuronal environment of the retina and choroid in exudative AMD. Separately, we observed enrichment of GO categories for regulation of cell killing, regulation of leukocyte mediated cytotoxicity, cell killing, and leukocyte mediated cytotoxicity; all consistent with alterations to signaling pathways of cytotoxic lymphocytes. We next identified the top highly enriched functional protein pathways in our data (Figure 2B). The top 5 pathways represented were: adrenaline and noradrenaline biosynthesis, dopamine receptor mediated signaling pathway, T-cell activation, interleukin signaling pathway, and the notch signaling pathway. VEGF and angiogenesis pathways were also found to be enriched in our data (despite no difference detected in actual VEGF levels), consistent with exudative AMD pathology and a compensatory angiogenic signal resulting from anti-VEGF therapy.^8^

**Figure 2.**
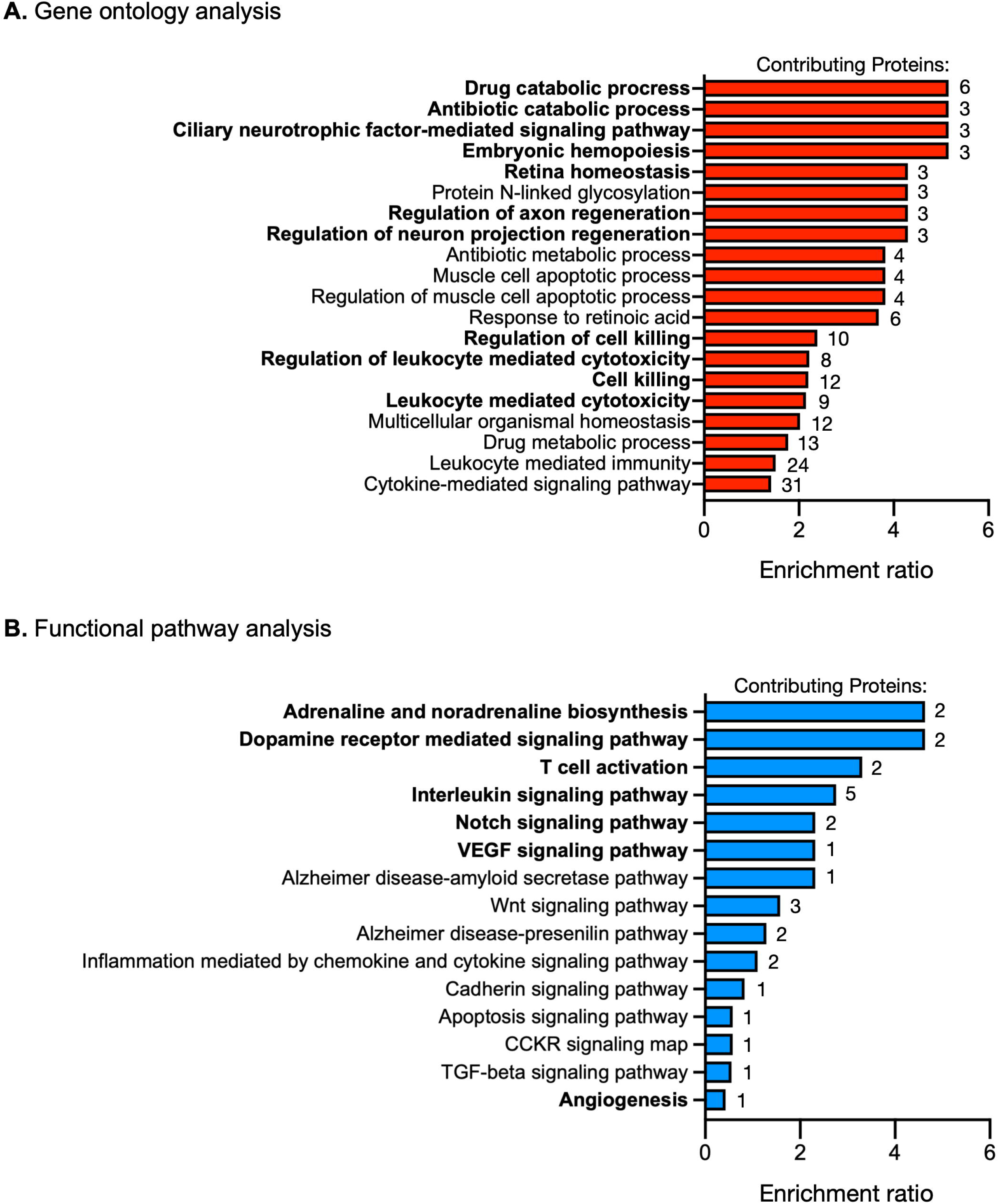
Gene ontology and PANTHER pathway analyses reveal key pathways in exudative AMD. Differentially expressed proteins (p<0.05) were entered into the WebGestalt online analysis tool. The number of genes contributing to the enrichment score from each gene set is listed to the right, beside each bar. **A)** Samples underwent Gene Ontology analysis and the categories with the highest enrichment ratios are shown. The top five upregulated gene categories in AMD samples compared with controls were: drug catabolic process, antibiotic catabolic process, ciliary neurotrophic factor-mediated signaling pathway, embryonic hemopoiesis, and retina homeostasis. **B)** Similarly, samples underwent PANTHER pathway analysis using WebGestalt and the pathways with the highest enrichment ratios are shown. The top five upregulated pathways in AMD samples compared with controls were: adrenaline and noradrenaline biosynthesis, dopamine receptor mediated signaling pathway, T-cell activation, interleukin signaling pathway, and Notch signaling pathway. Top pathways and those highlighted in the text are bolded.

### Drug repositioning analysis identifies four relevant drugs

Finally, we identified existing pharmacological treatments with relevance to this proteomic composition (Figure 3). CTLA4, an inhibitory cytotoxic T-cell receptor, was highly downregulated. Abatacept is a fusion protein of CTLA4 and IgG1, already approved for rheumatoid arthritis. Repositioning abatacept could competitively inhibit CD80/86 on antigen presenting cells and prevent co-stimulation of T-cells in exudative AMD. The upregulated IL-2R pathway could be targeted through the monoclonal antibody daclizumab, a disease-modifying treatment for multiple sclerosis, or small molecule inhibitors, such as sirolimus, an mTOR inhibitor used in transplant rejection. Finally, dupilumab, an anti-IL-4R antibody used in atopic diseases, could antagonize IL-4 driven Th2 polarization, which would reduce M2 macrophage differentiation during the delayed angiogenic phase of exudative AMD.

**Figure 3.**
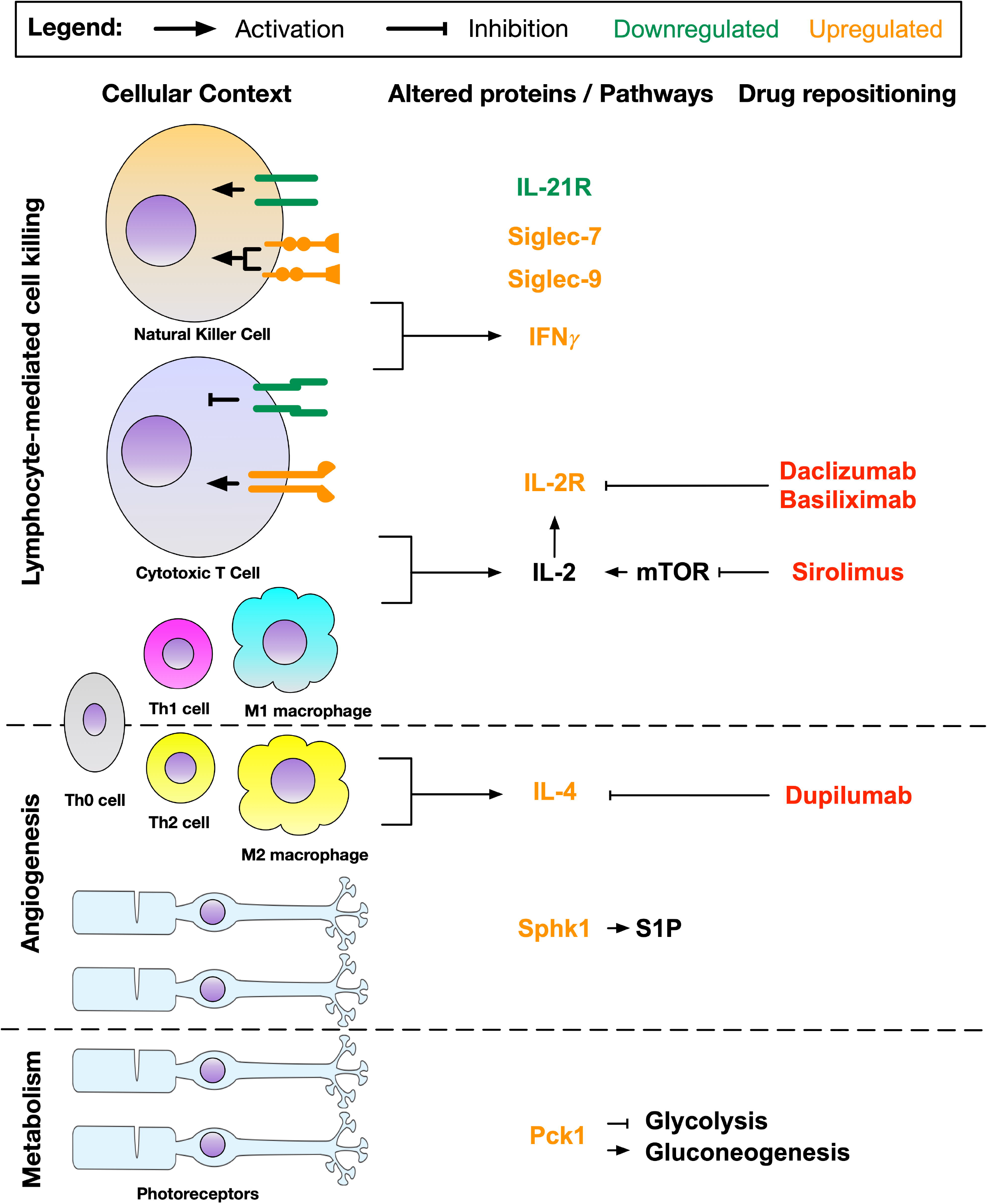
Drug repositioning model for exudative AMD therapy. Model of the pathogenesis of exudative AMD highlighting cells, molecules, and pathways implicated in our analysis. The three main pathways involved are: lymphocyte-mediated cell killing, angiogenesis, and cellular metabolism. Cytotoxicity due to upregulated cytokines (e.g. IL-2, IFNγ) and dysregulated receptors (e.g. SIGLEC-7/9, CTLA4) on NK cells, cytotoxic T cells, and M1 macrophages may initiate or exacerbate destruction of the blood-retina-barrier (BRB). Subsequently, angiogenesis and wound healing mediated by M2 macrophages, photoreceptors, and RPE (not shown) is represented through IL-4 and SPHK1 upregulation. Metabolic changes such as PCK1 upregulation may also contribute to an altered angiogenic state. Cytokines listed may also be released by other cells, but for simplicity, only key pathways are shown. Relevant therapeutics which have been approved for other diseases that may be repurposed for use in exudative AMD are indicated with yellow boxes and red text.

## DISCUSSION

While anti-VEGF therapy effectively suppresses neovascularization in most cases of exudative AMD, many patients continue to experience progressive retinal degeneration and incomplete visual recovery. This suggests that VEGF-independent mechanisms contribute to disease persistence. Here, we provide human vitreous proteomic evidence that cytotoxic immune signaling pathways remain active despite anti-VEGF treatment, supporting a model in which immune dysregulation represents a parallel and potentially targetable axis of disease.

Our data implicate dysregulation of cytotoxic lymphocyte signaling, including both natural killer (NK) cells and cytotoxic T lymphocytes, as a prominent feature of treated AMD. The concurrent downregulation of IL-21R and upregulation of SIGLEC-7/9 suggests a complex regulatory state in which cytotoxic immune cells exhibit altered activation thresholds rather than simple activation or suppression. SIGLEC family receptors function as inhibitory immune checkpoints, and their upregulation may reflect compensatory feedback to chronic immune activation.^26,27^ In parallel, reduced expression of CTLA4—an inhibitory receptor on T cells— suggests disinhibition of cytotoxic T-cell activity. Together, these findings indicate a dysregulated immune environment characterized by competing activating and inhibitory signals that may contribute to ongoing retinal injury. The observed NK cell–associated signals further support this interpretation. IL-21 promotes NK cell proliferation and maturation, whereas SIGLEC-7 inhibits NK-mediated cytotoxicity, suggesting disruption of a finely tuned regulatory balance.^28,29^ Although these findings implicate NK cells in AMD pathology following anti-VEGF treatment, it remains unclear whether they reflect changes in cell abundance or functional state. The concurrent upregulation of IFNγ supports enhanced cytotoxic activity,^30^ though additional studies are needed to resolve these dynamics.

Importantly, SIGLEC-7 upregulation was observed in both AMD and vitreous hemorrhage cohorts, raising the possibility of a broader role in vitreoretinal pathology. However, the lack of correlation with hemorrhage status argues against simple blood contamination as a confounder. Rather, SIGLEC-7 may represent a context-dependent marker of immune activation within the diseased vitreous. While not necessarily specific to AMD, its consistent dysregulation supports its relevance as a candidate biomarker and potential therapeutic target. Given the overlap between NK and CD8+ T-cell signaling markers, these findings likely reflect broader cytotoxic lymphocyte involvement.^31^ Downregulation of CTLA4 further supports increased cytotoxic T-cell activity, consistent with enriched T-cell activation pathways identified in our gene ontology analysis. Prior studies have similarly implicated cytotoxic lymphocytes in AMD, including associations with NK cell receptor haplotypes,^35^ detection of NK cells in subretinal membranes,^36^ and evidence that retinal pigment epithelial cells can suppress NK cell activity under normal conditions.^37^ Loss of this regulatory capacity may contribute to immune dysregulation in disease. Additionally, although SIGLEC proteins are most commonly associated with peripheral immune cells, their expression on microglia raises the possibility that these signals may also reflect resident immune populations within the retina.^38^

The identification of dysregulated immune pathways in this study provides a rationale for therapeutic strategies targeting cytotoxic lymphocyte signaling in AMD. Several clinically approved immunomodulatory agents—including abatacept, sirolimus, and dupilumab—intersect with pathways identified in our dataset. Notably, prior clinical studies have demonstrated that systemic sirolimus or daclizumab reduces anti-VEGF treatment burden by approximately 50% when used in combination therapy.^39^ These findings suggest that modulation of IL-2/IL-2R and related immune pathways may enhance therapeutic durability by addressing VEGF-independent disease mechanisms.

We observed limited concordance between this study and prior analyses of aqueous humor from AMD patients.^8^ This discrepancy likely reflects differences in sampling location, patient populations, and proteomic methodology. As vitreous sampling occurs in closer proximity to the site of pathology and avoids some of the limitations of anterior chamber sampling, the molecular signals identified here may more accurately reflect the disease microenvironment.

This study has several limitations. Multiplex antibody arrays are restricted to predefined targets, and thus do not capture the full proteomic landscape, including many immune and complement-related proteins. While C5a was included in our panel and did not differ between groups,^40^ broader complement pathway involvement cannot be excluded. Additionally, this study is correlative and does not establish causality. As with all discovery-stage proteomic studies, validation in independent cohorts and with orthogonal methods will be necessary.^10^ Future studies incorporating larger sample sizes, earlier disease stages, and treatment-naïve patients will be particularly informative, although acquisition of vitreous from untreated AMD patients remains challenging due to current treatment paradigms.^41^

In summary, vitreous proteomic analysis of anti-VEGF–treated AMD identifies persistent cytotoxic immune signaling alongside angiogenic and metabolic alterations. These findings support a model in which immune dysregulation contributes to incomplete therapeutic response and highlight cytotoxic lymphocyte pathways as potential targets for combination treatment strategies.

## Data Availability

All data produced in the present study are available upon reasonable request to the authors

## Author Contributions

Dr. Mahajan had full access to all the data in the study and takes responsibility for the integrity of the data and the accuracy of the data analysis. Study concept and design: M.A.T., S.H.T., A.G.B., V.B.M. Acquisition of data: M.A.T., G.V., V.B.M. Analysis and interpretation of data: M.A.T., B.N., G.V., V.B.M. Drafting of the manuscript: M.A.T., B.N., A.G.B., S.H.T., V.B.M. Critical revision of the manuscript for important intellectual content: A.G.B., V.B.M., S.H.T. Statistical analysis: M.A.T., J.Y., V.B.M. Obtained funding: V.B.M. Administrative, technical, and material support: A.G.B., S.H.T., V.B.M. Study supervision: V.B.M.

---

**Supplemental Figure 1.**
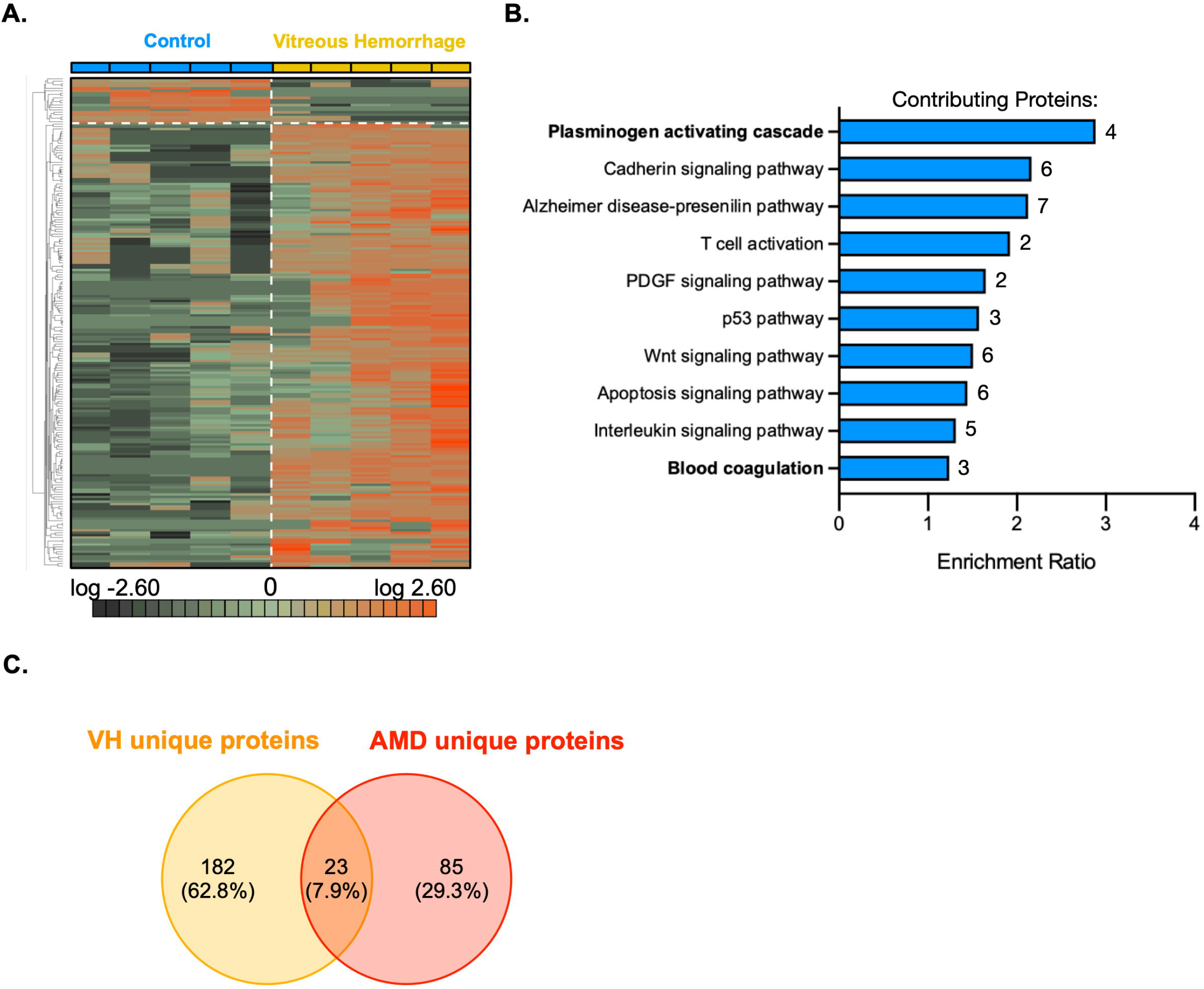
Vitreous hemorrhage proteomic signal differs from AMD. **A.** Five vitreous hemorrhage (VH) and five comparative control vitreous biopsies were used for proteomic analysis as in Figure 1. Protein levels (pg/ml) from the multiplex ELISA assay were analyzed by 1-way ANOVA and differentially-expressed proteins (p<0.05) were represented as a heat map. 205 differentially expressed proteins were identified in total, with 19 down-regulated in VH and 186 upregulated. **B.** PANTHER pathway analysis was performed on this dataset, as in Figure 2, to identify the most active protein pathways in VH compared to controls. The number of genes contributing to the enrichment score from each gene set is listed to the right beside each bar. **C.** A Venn diagram comparison of differentially expressed proteins identified in VH and AMO. 182 unique VH proteins were identified and 85 unique AMO proteins were identified, while 23 shared proteins were found to be differentially expressed in both conditions.

